# Benefits of a novel bimanual urethral catheterization technique for female patients: a nursing school experiment on skills, hygiene and duration

**DOI:** 10.1101/2025.04.24.25326249

**Authors:** M. Holl, A. Benkiser, L. Kloppenburg, U. Frey, U. Schneider

**Affiliations:** Fraunhofer Institute for Production and Automation Technology (IPA) Stuttgart and University of Stuttgart Institute of Industrial Manufacturing and Management (IFF) University of Stuttgart, Germany; Oberlinhaus Nursing School, Freudenstadt Germany

## Abstract

Urethral catheterization in female patients is a psychologically, ergonomically and hygienically demanding task. A student experiment in a nursing school on manikins could show that the new thigh and labia spreading device by Urs Schneider may improve comfort for patients and nurses, reduce duration and physical stress, and facilitate hygiene rule fulfillment subjectively for the catheterization process by having two free hands and a thigh and labia spreading device as a third hand. The students also had the impression that hygiene rules in catheterization are easier to fulfill with the new device.

## Background

Urethral catheterization is a relevant task for physicians and nurses in daily intermittent application on patients suffering from neurogenic bladder paralyses and when placing indwelling catheters for inpatients or in bedridden elderly people. Although catheterization of male patients can be demanding, catheterization in women can be very stressful (V. Geng et al.^i^): wide spreading of the thighs for visibility and access combined with manual opening of the inner labia of a woman is and - remains lifelong - a very intimate task. Patients report on stress and even on nursing staff, who cannot identify the urethral entrance. Catheterization goes with critically high rates of urinary tract infections (Brinkhof et al.^ii^). A new thigh and labia spreader invented by Urs Schneider intents to facilitate catheterization in women (U. Schneider^iii^). The thigh spreading angle is significantly reduced and the inner labia are opened and kept open by a third hand mechanism. This enables two-handed catheterization, facilitates the one-minute disinfection time with spread labia and it relieves the strain from the nurses back, by reducing the demand for static forward bending during the task.

Research is part of the new nursing curriculum in Germany. Oberlinhaus nursing school, Freudenstadt, Fraunhofer Institute IPA and University Stuttgart Institute IFF collaborate in care innovation and training. A joint experiment was planned during care classes and an experiment was executed on patient manikins. The team wanted to learn about duration of catheterization, potentials for better sticking to hygiene rules in a bimanual setting, psychology and efficiency of catheterization.

## Methodology

The aim of the present study is to compare the catheterization technique on female patients using the single-handed method (without devices) and the two-handed method (with device). The German educational check list for urethral catheterization of an indwelling catheter in female patients was used (S. Urban^iv^). The duration from labia spreading (manual versus device based) until checking whether the catheter is sufficiently blocked was measured and compared. The students developed the following questions:

1. In how far do you think that hygiene rules are easier fulfilled with two free hands relative to the standard single hand technique?
2. Is the two-handed procedure perceived as being more comfortable for patients and nurses?
3. Is the two-handed procedure perceived as being more effective for patients and nurses?
4. Is the bimanual procedure less strenuous for nurses than the single-handed procedure?

## Results

16 nurses in training (15 female and 1 male) with an average age of 26.81+- 9.17 years performed the catheterization method of sterile placement of a urinary catheter according to the Pflegias textbook in randomized order once single-handed and once with the device on a manikin.

The subjective effort was assessed using the BORG scale (from very, very easy 6 to very, very strenuous 20), the duration of the procedure using video cameras, as well as the hygiene management and well-being of the nurse and patient. The average time from “open labia” to “check catheter blockage” was 2.62 min without the new device and 2.19 min with the new device as in Figure 1.

**Figure 1.**
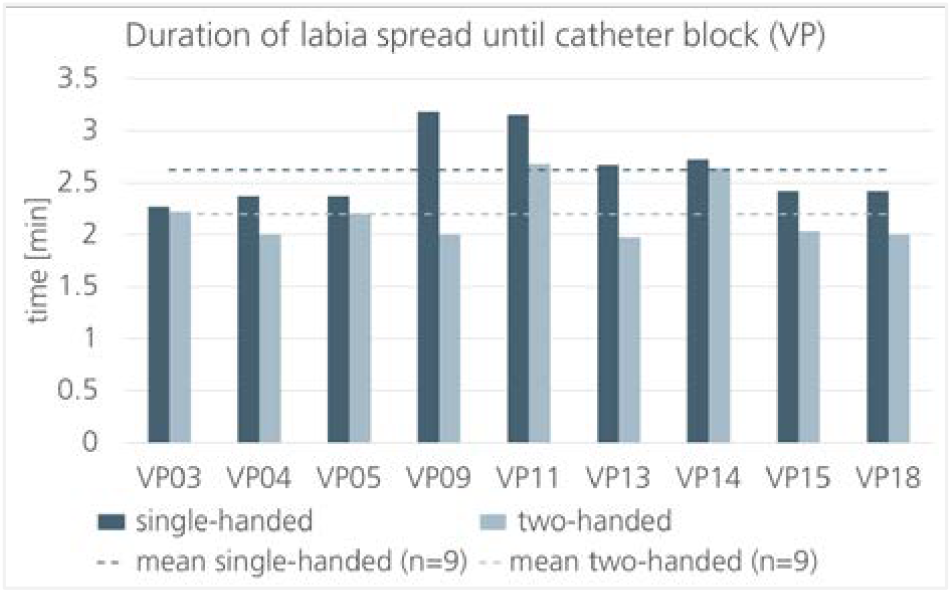
Compared duration of catheterization process from labia spreading until catheter block check single-handed (without device) compared to two-handed (using the new device), n=9.

The subjective effort was lower for most nurses with the new device (Figure 2). 6 out of 9 nurses perceived catheterization without the aid as strenuous, while it was perceived as easier with the device. During the evaluation of the questionnaire and the video data, seven test subjects had to be excluded because they did not sufficiently adhere to the protocols. The students had the impressions that hygiene rules in catheterization are easier to fulfill with the new device.

**Figure 2.**
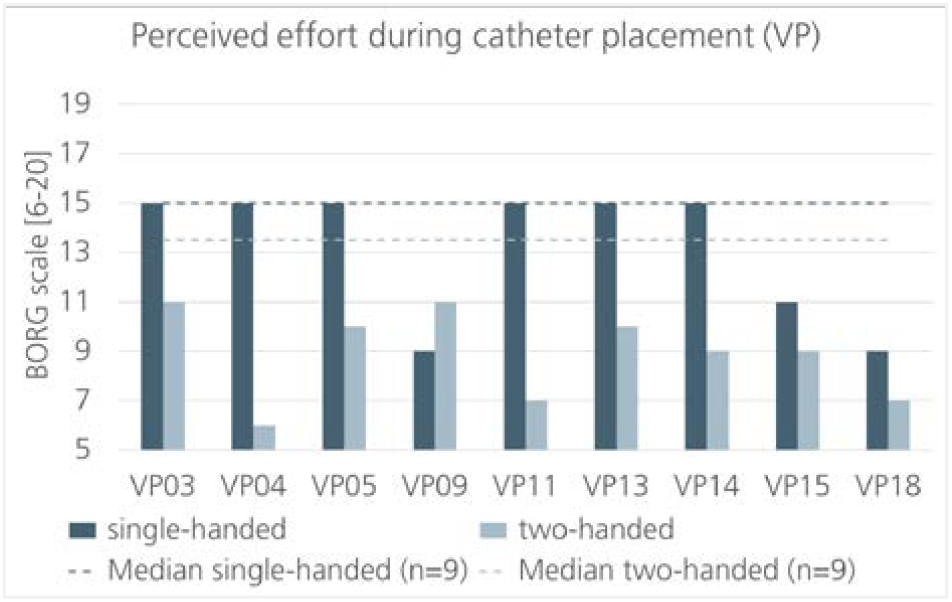
Compared effort of complete catheterization process single-handed compared to two-handed (using the new device), n=9.

### Interpretation

The study shows that the new thigh and labia spreader by Urs Schneider may make external catheterization easier, slightly faster and more comfortable for both nurses and patients. The students also had the impressions that hygiene rules in catheterization are easier to fulfill with the new device. Further studies with more nursing students and with experienced nurses have to follow.

## Data Availability

All data produced in the present study are available upon reasonable request to the authors

